# A Composite Index to Measure Family Planning Performance with Application to India

**DOI:** 10.1101/2020.12.16.20248368

**Authors:** Aalok Ranjan Chaurasia

## Abstract

This paper develops a composite family planning performance index that takes into account the met demand for modern family planning methods and the structure of the met demand or the method mix. Application of the index to India suggests that, although, there has been an improvement in India’s family planning performance, yet it has remained poor throughout the period 1992 through 2016 and there are significant variation in the performance across states/Union Territories and across districts. The classification modelling exercise reveals that districts in India can be grouped into eight clusters in terms of family planning performance and both met demand for modern family planning methods and the structure of the met demand are essentially different in different clusters. The paper emphasises that family planning in the country must be promoted as a development strategy rather than an intervention to limit births.

## Family Planning Performance in India: 1992-2016

### Introduction

The performance of family planning efforts is conventionally measured in terms of the contraceptive prevalence rate (CPR). The popularity of CPR to measure family planning performance rests with its strong inverse relationship with the total fertility rate (TFR) based on cross-country data (Bongaarts, 1978; Bongaarts and Potter, 1983; Ross and Mauldin, 1996; Jain, 1997; Tsui, 2001; Stover, 1998; United Nations, 2020). There are, however, studies that show inconsistency between CPR and TFR in many countries (United Nations, 2020). Using state level data from India, Srinivasan (1988) has argued that as one goes down the level of aggregation, variation in family planning use explains less and less of the variation in TFR. Chaurasia (2004), using development block level data from Madhya Pradesh, India has observed that variation in the use of family planning methods among currently married reproductive age couples explains only around 20 per cent of the variation in total marital fertility rate. Chaurasia (2000) has also observed that the number of children ever born is positively related with family planning use at the individual level.

The CPR, defined as the proportion of currently married women of reproductive age using a family planning method, however, is not an appropriate indicator to measure family planning performance. CPR is essentially a ratio, not a rate or incidence of family planning use. It is, essentially a crude measure that does not take into account the structure of family use or the distribution of family planning users by different family planning methods. The upper limit of CPR is also difficult to establish. It is well-known that a substantial proportion of currently married reproductive age women do not practice family planning method because they either want child or are pregnant or the woman or her partner is sterile and this proportion is not constant but varies widely across different population groups. CPR is also insensitive to the fact that family planning needs of a couple vary by different phases of the family building process and are conditioned by such factors as personal circumstances, individual knowledge and changing childbearing preferences. Availability and accessibility of different family planning methods and their effectiveness also affect use of different family planning methods. It has, therefore, been emphasised that any measure of family planning performance must also take into account the range and types of family planning methods being used (United Nations, 2019). The CPR, neither captures the met demand for family planning nor the structure or the composition of the demand.

The above considerations suggest that family planning performance should be measured on a two-dimensional space. One dimension of the performance space should reflect the met demand for family planning while the other dimension should reflect the structure of the met demand or the method mix. The method mix reflects the extent up to which family planning efforts are able to meet the diverse family planning needs of the people and, therefore, is an important component of family planning performance. The CPR completely overlooks the structure of the met demand for family planning and, therefore, depicts a biased picture of family planning performance.

In this paper we construct a composite index of family planning performance that takes into account both the met demand for family planning and the structure of the met demand or the method mix. The performance index so constructed has been used to measure the level, trend and spatial variations in the family planning performance in India. Organised family planning efforts in India date back to 1952 when the country launched world’s first official family planning programme. In its initial phase, main appeal of family planning was health and welfare of the family, especially children and women. (Chaurasia and Singh, 2014). Subsequently, focus of official family planning efforts shifted towards birth limitation to reduce fertility and control population growth. In its initial phase, the only source of family planning related information was the programme service statistics of the official family planning programme and indicators like equivalent sterilisation and couples effectively protected were used to measure family planning performance (Chaurasia, 1985; Government of India, 1990). The first nationally representative family planning survey was carried out in 1970 which revealed that about 14 per cent of the currently married women aged 15-44 years or their husband were using a family planning method, while less than 10 per cent were using a modern method (Operations Research Group, 1970). The second all India survey, carried out in 1980, revealed that the proportion of currently married women aged 15-44 years or their husband using a family planning method had increased to more than 35 per cent while more than 28 per cent were using a modern method (Khan and Prasad, 1980). In 1992, the National Family Health Survey (NFHS) Programme was launched and NFHS 1992-93, revealed a CPR of 40 per cent while the prevalence of modern methods (mCPR) was around 36 per cent (International Institute for Population Sciences, 1995). The NFHS 1998-99 revealed that the CPR had increased to almost 45 per cent (International Institute for Population Sciences and ORC Macro, 2000) while NFHS 2005-06 estimated a CPR of 55 per cent (International Institute for Population Sciences and Macro International, 2007). However, NFHS 2015-16, reported a decrease in CPR to 53 per cent while mCPR stagnated at less than 48 per cent (International Institute for Population Sciences and ICF, 2017). There has, however, never been any attempt to measure performance in terms of met demand and the structure of the met demand.

The paper is organised as follows. The next section of the paper constructs the composite family planning performance index on the basis of the two dimensional performance space discussed above. In section three of the paper, family planning performance in India and in its constituent states and Union Territories has been analysed during the 25 years period between 1992 and 2016. Inter-district variation in family planning performance, based on the composite family planning performance index are analysed in section four. The fifth section of the paper presents findings of the classification modelling exercise that classifies districts in terms of family planning performance on the basis of the performance on the basis of the met demand for family planning and structure of the met demand. Findings of the performance measurement analysis are discussed in section six in the context of meeting the diverse family planning needs of the people. The last section of the paper summarises the main findings of the analysis and discusses policy and programme options for improving family planning performance in India, especially, at the district level.

### Family Planning Performance Index

The underlying principles of family planning efforts in the context of meeting the diverse family planning needs of the people are informed choice and provision of a range of family planning methods to meet the needs of potential family planning users. If *p* denotes the family planning performance index, *c* denotes the index of met demand for family planning and *q* denotes the index of the structure of the met demand, then the index *p* can be related to *c* and *q* through an appropriate aggregation function *f*. In other words

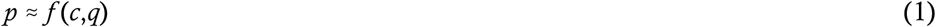

The above performance measurement framework can be operationalised by specifying the aggregation function *f* and defining indexes *c* and *q*. The simplest of the aggregation function is the simple arithmetic mean. However, an undesirable feature of the simple arithmetic mean is its additive compensability (OECD, 2000). The other alternative is to use power mean or generalised mean (Bullen, 2003; Stanislav, 2009). Anand and Sen (1997) have used power mean to construct a multidimensional poverty index. Chaurasia (2018) has used weighted power mean to construct a development index for Indian villages. Using the weighted power mean as the aggregation function, the family planning performance index, *p*, may be constructed as:

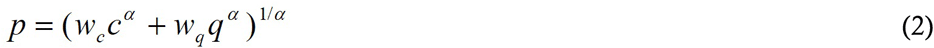

where *w*_*c*_ is the weight assigned to the index, *c*, and *w*_*q*_ is the weight assigned to the index, *q*. The parameter α, in equation (2) has an important impact on the index *p*. When α=1 and *w*_*c*_ =*w*_*q*_ =1, *p* is nothing but the simple arithmetic mean of *c* and *q*. When α<1, relatively more weight is given to that index which has a lower value then the index which has a higher value. There is, however, an inescapable arbitrariness in the selection of α. Theoretically, α can be any value <1. When α=1/3, the importance of the index having a lower value is four times the importance of the index having higher value in determining the performance index. When α=1/5, the relative importance of the index having lower value is 16 times the importance of the index having higher value. For α<1/5, the difference turns out to be even wider. However, as performance improves, the difference narrows down and when the two indexes are the same, the difference in the relative importance becomes zero. Assigning unequal weights to contributed indexes is justified on the ground that it provides impetus to improve that component in which the performance is poor.

Choosing α=1/3 and assigning equal weights to indexes *c* and *q*, the family planning performance index, *p*, is defined as 

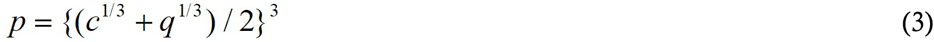

An important requirement for the performance index, *p*, defined by equation (3), is that the variation in the index, *c*, must be independent of the variation in the index, *q*, but both *c* and *q* must covary with the index, *p*. Mutual independence of indexes *c* and *q* is important as it ensures that improvement in both met demand for family planning and the structure of the met demand is necessary to improve family planning performance in the context of meeting the diverse family planning needs of the people.

The above family planning performance measurement framework requires constructing the index *c* of the met demand for family planning and the index *q* of method mix. The met demand for family planning can be classified further into met demand for modern spacing methods and met demand for permanent methods. The index of the met demand for modern spacing methods, *c*_*s*_, may be constructed as: 

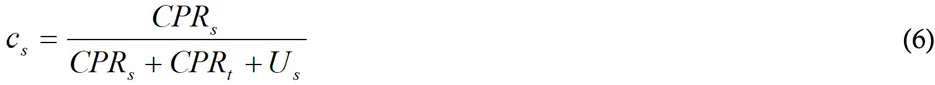

where *CPR*_*s*_ is the prevalence of modern spacing methods, *CPR*_*t*_ is the prevalence of traditional methods and *U*_*s*_ is the unmet need for spacing methods. It is assumed that the use of traditional methods actually reflects the unmet demand for modern spacing methods.

On the other hand, the met demand for permanent methods, *c*_*p*_, may be constructed as 

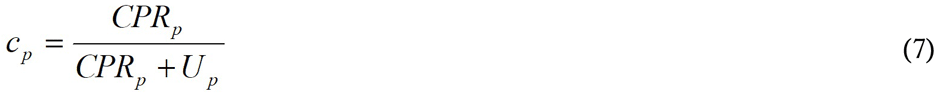

where *CPR*_*p*_ is the prevalence of permanent methods of family planning and *U*_*p*_ is the unmet need for terminal methods.

Based on the indexes *c*_*s*_ and *c*_*p*,_ the index *c* reflecting the met demand for modern family planning methods is constructed as 

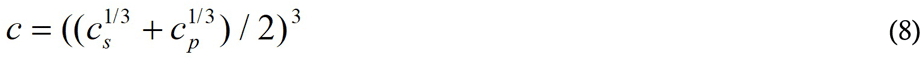

Since, both *c*_*s*_ and *c*_*p*_ vary between 0 and 1, *c* also varies between 0 and 1. The higher the index *c*, the higher the met demand for modern family planning methods.

On the other hand, index *q* reflecting the structure of the met demand or the method mix may be constructed using the concept of the dominance of one family planning method over others. Using this concept and following Chaurasia (2020), the index *q* reflecting the structure of the met demand or the method mix is constructed as 

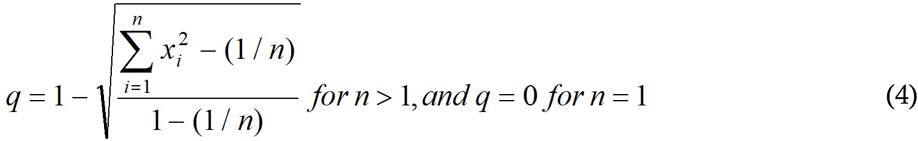

where *x*_*i*_ is the proportionate prevalence rate of the method *i* and *n* is the total number of modern family planning methods available. The index *q* is independent of the number of modern family planning methods available and ranges from 0 to 1. The higher the value of *q* the more uniform is the distribution of family planning users by different family planning methods.

It is obvious that the family planning performance index, *p*, varies between 0 and 1 and the higher the index *p*, the better the family planning performance and vice versa. Based on the index *p*, family planning performance may be classified as very poor if *p*<0.300; poor if 0.300≤*p*<0.550; average if 0.550≤*p*<0.750; good if 0.750≤*p*<0.900; and very good if *p*≥0.900. The index *p* along with indexes *c* and *q* constitutes a comprehensive family planning performance measurement framework. We use this framework to measure family planning performance in India.

### Family Planning Performance in India

Estimates of the prevalence of modern family planning methods along with the unmet need for modern spacing methods and permanent methods of family planning in India are available from different rounds of the National Family Health Survey (NFHS) carried out in 1992-93, 1998-99, 2005-06 and 2015-16 (Table 1). The first three rounds of NFHS provided method-specific prevalence rates for the country and for its constituent states/Union Territories. The fourth round of NFHS, however, also provided estimates of method-specific prevalence rates for 640 districts of the country as they existed at the time of the 2011 population census. Based on these estimates, indexes *c*_*s*_, *c*_*p*_, *c, q* and the performance index *p* have been calculated for the country and for its constituent states/Union Territories and 640 districts.

**Table 1.** Family planning performance in India, 1992-93 through 2015-16

| Particulars | Period |  |  |  |
| --- | --- | --- | --- | --- |
|  | 1992-93 | 1998-99 | 2005-06 | 2015-16 |
| *CPR* | 40.7 | 48.2 | 56.3 | 53.5 |
| *CPR<sub>m</sub>* | 36.5 | 42.8 | 48.5 | 47.8 |
| Method-specific prevalence |  |  |  |  |
| Female sterilisation | 27.4 | 34.1 | 37.3 | 36.0 |
| Male sterilisation | 3.5 | 1.9 | 1.0 | 0.3 |
| IUD | 1.9 | 1.6 | 1.7 | 1.5 |
| Oral pill | 1.2 | 2.1 | 3.1 | 4.1 |
| Condom | 2.4 | 3.1 | 5.2 | 5.6 |
| Other modern methods | 0.1 | 0.0 | 0.2 | 0.3 |
| Traditional methods | 4.2 | 5.4 | 7.8 | 5.7 |
| Unmet need for spacing | 12.2 | 8.3 | 6.1 | 5.6 |
| Unmet need for limiting | 8.1 | 7.8 | 7.8 | 7.2 |
| Quality index (*q*) | 0.295 | 0.242 | 0.272 | 0.288 |
| Coverage index (*c<sub>s</sub>*) | 0.257 | 0.338 | 0.429 | 0.502 |
| Coverage index (*c<sub>p</sub>*) | 0.792 | 0.822 | 0.831 | 0.834 |
| Coverage index © | 0.475 | 0.545 | 0.608 | 0.654 |
| Performance index (*p*) | 0.378 | 0.373 | 0.417 | 0.446 |

Source: Author's calculations

The performance index *p* in India is estimated to have increased during the 25 years between 1992-93 and 2015-16 but the family planning performance in the country may be rated as poor throughout this period. The met demand for modern family planning methods increased throughout this period but the method mix has remained highly skewed. Moreover, there remains a big gap between the met demand for modern spacing methods and the met demand for permanent methods, although the gap has narrowed over time.

Among states/Union Territories, family planning performance has been found to be relatively the best in Sikkim but the poorest in Andhra Pradesh during 2015-16 (Table 2). There is no state/Union Territory in the country where family planning performance may be rated as good or very good whereas, in 28 of the 36 states/Union Territories, family planning performance remains either poor or very poor. The inter-state/Union Territory variation in both capacity index and coverage index has contributed to the inter-state/Union Territory variation in family planning performance as reflected by the index *p*. There is, however, no state where both capacity index and coverage index are rated as very good. By contrast, the capacity index is rated as very poor in 17 states whereas the coverage index is rated as poor in only one state during 2015-16.

**Table 2.** Indicators of family planning performance in states/Union Territories of India

| State/Union Territory | 1992-93 | 1998-99 | 2005-06 | 2015-16 |
| --- | --- | --- | --- | --- |
| | Index $p$ | | | |
| Andaman and Nicobar Islands | na | na | na | 0.376 |
| Andhra Pradesh & Telangana | 0.314 | 0.279 | 0.235 | 0.166 |
| Arunachal Pradesh | 0.469 | 0.46 | 0.505 | 0.525 |
| Assam | 0.371 | 0.448 | 0.459 | 0.492 |
| Bihar & Jharkhand | 0.285 | 0.235 | 0.315 | 0.264 |
| Chandigarh | na | na | na | 0.628 |
| Dadra & Nagar Haveli | na | na | na | 0.339 |
| Daman and Diu | na | na | na | 0.35 |
| Delhi | 0.659 | 0.639 | 0.635 | 0.606 |
| Goa | 0.365 | 0.364 | 0.439 | 0.442 |
| Gujarat | 0.385 | 0.402 | 0.450 | 0.408 |
| Haryana | 0.478 | 0.480 | 0.562 | 0.577 |
| Himachal Pradesh | 0.520 | 0.455 | 0.569 | 0.51 |
| Jammu & Kashmir | 0.474 | 0.468 | 0.555 | 0.589 |
| Karnataka | 0.299 | 0.273 | 0.290 | 0.223 |
| Kerala | 0.384 | 0.306 | 0.346 | 0.264 |
| Lakshadweep | na | na | na | 0.352 |
| Madhya Pradesh & Chhattisgarh | 0.364 | 0.334 | 0.374 | 0.365 |
| Maharashtra | 0.422 | 0.405 | 0.460 | 0.437 |
| Manipur | 0.508 | 0.453 | 0.522 | 0.395 |
| Meghalaya | 0.403 | 0.422 | 0.415 | 0.495 |
| Mizoram | 0.374 | 0.425 | 0.488 | 0.555 |
| Nagaland | 0.430 | 0.454 | 0.483 | 0.499 |
| Odisha | 0.294 | 0.313 | 0.439 | 0.507 |
| Puducherry | na | na | na | 0.262 |
| Punjab | 0.555 | 0.604 | 0.615 | 0.613 |
| Rajasthan | 0.303 | 0.361 | 0.426 | 0.443 |
| Sikkim | na | 0.519 | 0.626 | 0.678 |
| Tamil Nadu | 0.341 | 0.288 | 0.294 | 0.250 |
| Tripura | 0.44 | 0.490 | 0.534 | 0.484 |
| Uttar Pradesh & Uttarakhand | 0.383 | 0.375 | 0.443 | 0.48 |
| West Bengal | 0.383 | 0.459 | 0.488 | 0.595 |
| Andhra Pradesh | na | na | na | 0.133 |
| Bihar | na | na | 0.295 | 0.236 |
| Chhattisgarh | na | na | 0.368 | 0.37 |
| Jharkhand | na | na | 0.369 | 0.339 |
| Madhya Pradesh | na | na | 0.373 | 0.363 |
| Telangana | na | na | na | 0.207 |
| Uttar Pradesh | na | na | 0.433 | 0.476 |
| Uttarakhand | na | na | 0.591 | 0.57 |
| Index $q$ | | | | |
| Andaman and Nicobar Islands | na | na | na | 0.205 |
| Andhra Pradesh and Telangana | 0.206 | 0.124 | 0.073 | 0.034 |
| Arunachal Pradesh | 0.505 | 0.406 | 0.443 | 0.539 |
| Assam | 0.449 | 0.453 | 0.486 | 0.439 |
| Bihar and Jharkhand | 0.236 | 0.170 | 0.229 | 0.153 |
| Chandigarh | na | na | na | 0.521 |
| Dadra and Nagar Haveli | na | na | na | 0.191 |
| Daman and Diu | na | na | na | 0.224 |
| Delhi | 0.597 | 0.552 | 0.538 | 0.529 |
| Goa | 0.26 | 0.261 | 0.343 | 0.353 |
| Gujarat | 0.238 | 0.230 | 0.283 | 0.257 |
| Haryana | 0.384 | 0.319 | 0.388 | 0.403 |
| Himachal Pradesh | 0.438 | 0.302 | 0.356 | 0.366 |
| Jammu & Kashmir | 0.420 | 0.387 | 0.479 | 0.518 |
| Karnataka | 0.159 | 0.105 | 0.097 | 0.063 |
| Kerala | 0.271 | 0.161 | 0.186 | 0.106 |
| Lakshadweep | na | na | na | 0.335 |
| Madhya Pradesh and Chhattisgarh | 0.298 | 0.192 | 0.194 | 0.176 |
| Maharashtra | 0.28 | 0.226 | 0.251 | 0.220 |
| Manipur | 0.587 | 0.483 | 0.679 | 0.657 |
| Meghalaya | 0.431 | 0.582 | 0.522 | 0.501 |
| Mizoram | 0.186 | 0.243 | 0.316 | 0.476 |
| Nagaland | 0.583 | 0.499 | 0.594 | 0.566 |
| Odisha | 0.217 | 0.188 | 0.297 | 0.405 |
| Puducherry | na | na | na | 0.074 |
| Punjab | 0.442 | 0.495 | 0.482 | 0.456 |
| Rajasthan | 0.214 | 0.227 | 0.268 | 0.272 |
| Sikkim | na | 0.508 | 0.628 | 0.686 |
| Tamil Nadu | 0.199 | 0.121 | 0.099 | 0.072 |
| Tripura | 0.464 | 0.407 | 0.476 | 0.382 |
| Uttar Pradesh and Uttarakhand | 0.425 | 0.366 | 0.429 | 0.448 |
| West Bengal | 0.346 | 0.366 | 0.388 | 0.472 |
| Andhra Pradesh | na | na | na | 0.019 |
| Bihar | na | na | 0.209 | 0.133 |
| Chhattisgarh | na | na | 0.203 | 0.181 |
| Jharkhand | na | na | 0.288 | 0.202 |
| Madhya Pradesh | na | na | 0.19 | 0.175 |
| Telangana | na | na | na | 0.059 |
| Uttar Pradesh | na | na | 0.427 | 0.45 |
| Uttarakhand | na | na | 0.446 | 0.448 |
| Index *c* |  |  |  |  |
| Andaman and Nicobar Islands | na | na | na | 0.623 |
| Andhra Pradesh and Telangana | 0.454 | 0.529 | 0.545 | 0.469 |
| Arunachal Pradesh | 0.434 | 0.519 | 0.572 | 0.510 |
| Assam | 0.303 | 0.444 | 0.433 | 0.548 |
| Bihar and Jharkhand | 0.34 | 0.315 | 0.420 | 0.419 |
| Chandigarh | na | na | na | 0.749 |
| Dadra and Nagar Haveli | na | na | na | 0.547 |
| Daman and Diu | na | na | na | 0.516 |
| Delhi | 0.724 | 0.734 | 0.743 | 0.690 |
| Goa | 0.494 | 0.491 | 0.551 | 0.546 |
| Gujarat | 0.584 | 0.645 | 0.674 | 0.609 |
| Haryana | 0.586 | 0.687 | 0.782 | 0.795 |
| Himachal Pradesh | 0.611 | 0.652 | 0.854 | 0.688 |
| Jammu & Kashmir | 0.532 | 0.560 | 0.639 | 0.667 |
| Karnataka | 0.503 | 0.562 | 0.645 | 0.542 |
| Kerala | 0.526 | 0.520 | 0.578 | 0.532 |
| Lakshadweep | na | na | na | 0.368 |
| Madhya Pradesh and Chhattisgarh | 0.439 | 0.532 | 0.640 | 0.655 |
| Maharashtra | 0.606 | 0.661 | 0.761 | 0.763 |
| Manipur | 0.436 | 0.424 | 0.391 | 0.214 |
| Meghalaya | 0.376 | 0.295 | 0.324 | 0.489 |
| Mizoram | 0.660 | 0.682 | 0.714 | 0.641 |
| Nagaland | 0.306 | 0.411 | 0.387 | 0.438 |
| Odisha | 0.387 | 0.483 | 0.619 | 0.625 |
| Puducherry | na | na | na | 0.639 |
| Punjab | 0.686 | 0.727 | 0.770 | 0.802 |
| Rajasthan | 0.415 | 0.538 | 0.638 | 0.673 |
| Sikkim | na | 0.529 | 0.624 | 0.669 |
| Tamil Nadu | 0.539 | 0.566 | 0.652 | 0.599 |
| Tripura | 0.417 | 0.583 | 0.596 | 0.602 |
| Uttar Pradesh and Uttarakhand | 0.344 | 0.384 | 0.456 | 0.513 |
| West Bengal | 0.423 | 0.566 | 0.605 | 0.738 |
| Andhra Pradesh | na | na | na | 0.427 |
| Bihar | na | na | 0.401 | 0.38 |
| Chhattisgarh | na | na | 0.605 | 0.659 |
| Jharkhand | na | na | 0.464 | 0.527 |
| Madhya Pradesh | na | na | 0.648 | 0.655 |
| Telangana | na | na | na | 0.501 |
| Uttar Pradesh | na | na | 0.439 | 0.503 |
| Uttarakhand | na | na | 0.764 | 0.713 |
| Index $c_s$ | | | | |
| Andaman and Nicobar Islands | na | na | na | 0.444 |
| Andhra Pradesh and Telangana | 0.18 | 0.253 | 0.267 | 0.182 |
| Arunachal Pradesh | 0.306 | 0.381 | 0.510 | 0.464 |
| Assam | 0.135 | 0.304 | 0.298 | 0.564 |
| Bihar and Jharkhand | 0.149 | 0.134 | 0.255 | 0.237 |
| Chandigarh | na | na | na | 0.673 |
| Dadra and Nagar Haveli | na | na | na | 0.365 |
| Daman and Diu | na | na | na | 0.339 |
| Delhi | 0.701 | 0.681 | 0.709 | 0.726 |
| Goa | 0.277 | 0.304 | 0.379 | 0.462 |
| Gujarat | 0.360 | 0.430 | 0.483 | 0.475 |
| Haryana | 0.397 | 0.512 | 0.711 | 0.719 |
| Himachal Pradesh | 0.394 | 0.447 | 0.801 | 0.610 |
| Jammu & Kashmir | 0.346 | 0.437 | 0.546 | 0.557 |
| Karnataka | 0.255 | 0.303 | 0.426 | 0.286 |
| Kerala | 0.264 | 0.260 | 0.328 | 0.284 |
| Lakshadweep | na | na | na | 0.158 |
| Madhya Pradesh and Chhattisgarh | 0.202 | 0.315 | 0.453 | 0.477 |
| Maharashtra | 0.393 | 0.461 | 0.622 | 0.637 |
| Manipur | 0.307 | 0.283 | 0.334 | 0.287 |
| Meghalaya | 0.168 | 0.246 | 0.236 | 0.47 |
| Mizoram | 0.437 | 0.488 | 0.570 | 0.589 |
| Nagaland | 0.291 | 0.329 | 0.424 | 0.422 |
| Odisha | 0.158 | 0.247 | 0.180 | 0.506 |
| Puducherry | na | na | na | 0.409 |
| Punjab | 0.551 | 0.601 | 0.707 | 0.705 |
| Rajasthan | 0.192 | 0.354 | 0.495 | 0.514 |
| Sikkim | na | 0.427 | 0.610 | 0.720 |
| Tamil Nadu | 0.309 | 0.339 | 0.455 | 0.372 |
| Tripura | 0.222 | 0.460 | 0.522 | 0.532 |
| Uttar Pradesh and Uttarakhand | 0.222 | 0.269 | 0.359 | 0.422 |
| West Bengal | 0.189 | 0.353 | 0.404 | 0.620 |
| Andhra Pradesh | na | na | na | 0.135 |
| Bihar | na | na | 0.228 | 0.203 |
| Chhattisgarh | na | na | 0.389 | 0.472 |
| Jharkhand | na | na | 0.323 | 0.343 |
| Madhya Pradesh | na | na | 0.468 | 0.479 |
| Telangana | na | na | na | 0.226 |
| Uttar Pradesh | na | na | 0.341 | 0.410 |
| Uttarakhand | na | na | 0.725 | 0.695 |
| Index $c_p$ | | | | |
| Andaman and Nicobar Islands | na | na | na | 0.844 |
| Andhra Pradesh and Telangana | 0.921 | 0.955 | 0.971 | 0.964 |
| Arunachal Pradesh | 0.594 | 0.688 | 0.638 | 0.56 |
| Assam | 0.573 | 0.621 | 0.603 | 0.533 |
| Bihar and Jharkhand | 0.649 | 0.612 | 0.644 | 0.674 |
| Chandigarh | na | na | na | 0.830 |
| Dadra and Nagar Haveli | na | na | na | 0.783 |
| Daman and Diu | na | na | na | 0.746 |
| Delhi | 0.748 | 0.790 | 0.778 | 0.656 |
| Goa | 0.803 | 0.742 | 0.769 | 0.639 |
| Gujarat | 0.887 | 0.923 | 0.908 | 0.766 |
| Haryana | 0.827 | 0.897 | 0.859 | 0.876 |
| Himachal Pradesh | 0.895 | 0.913 | 0.910 | 0.772 |
| Jammu & Kashmir | 0.775 | 0.706 | 0.741 | 0.789 |
| Karnataka | 0.873 | 0.939 | 0.928 | 0.917 |
| Kerala | 0.922 | 0.912 | 0.931 | 0.895 |
| Lakshadweep | na | na | na | 0.713 |
| Madhya Pradesh and Chhattisgarh | 0.813 | 0.831 | 0.874 | 0.874 |
| Maharashtra | 0.886 | 0.911 | 0.919 | 0.904 |
| Manipur | 0.597 | 0.605 | 0.453 | 0.155 |
| Meghalaya | 0.709 | 0.349 | 0.432 | 0.508 |
| Mizoram | 0.949 | 0.923 | 0.881 | 0.696 |
| Nagaland | 0.322 | 0.506 | 0.352 | 0.455 |
| Odisha | 0.771 | 0.834 | 0.782 | 0.761 |
| Puducherry | na | na | na | 0.943 |
| Punjab | 0.842 | 0.870 | 0.838 | 0.907 |
| Rajasthan | 0.766 | 0.778 | 0.806 | 0.861 |
| Sikkim | na | 0.648 | 0.638 | 0.621 |
| Tamil Nadu | 0.861 | 0.878 | 0.899 | 0.903 |
| Tripura | 0.702 | 0.726 | 0.675 | 0.678 |
| Uttar Pradesh and Uttarakhand | 0.504 | 0.529 | 0.569 | 0.616 |
| West Bengal | 0.796 | 0.849 | 0.864 | 0.87 |
| Andhra Pradesh | na | na | na | 0.979 |
| Bihar | na | na | 0.644 | 0.639 |
| Chhattisgarh | na | na | 0.889 | 0.89 |
| Jharkhand | na | na | 0.642 | 0.769 |
| Madhya Pradesh | na | na | 0.870 | 0.87 |
| Telangana | na | na | na | 0.939 |
| Uttar Pradesh | na | na | 0.554 | 0.608 |
| Uttarakhand | na | na | 0.805 | 0.732 |
| Source: | Author's calculations |  |  |  |
| Remarks | na: not available |  |  |  |

The trend in family planning performance has also varied across states/Union Territories. There are 25 states including undivided states of Andhra Pradesh, Bihar, Madhya Pradesh and Uttar Pradesh for which estimates of the prevalence rate of different modern family planning methods are available for both 1992-93 and 2015-16. Out of these 25 states, the performance index *p* decreased in 8 states whereas there are only 11 states, where both capacity index and coverage index have increased between 1992-93 and 2015-16. The undivided Andhra Pradesh, comprising of present Andhra Pradesh and Telangana, is the only state where both capacity index and coverage index decreased during 1992-16. The family planning coverage index decreased in Manipur and Mizoram also.

District level estimates of method specific prevalence rate, obtained from NFHS 2015-16, permit assessing family planning performance at the district level. The evidence available from NFHS 2015-16 suggests that there is no district where family planning performance may be rated as good or very good whereas in more than 85 per cent districts of the country, the performance may is either poor or very poor (Table 3). This leaves only around 15 per cent districts where performance may be termed as average. The Inter-district variation in the capacity index is contrastingly different from that in coverage index. There is no district where capacity index is good or very good whereas coverage index is very good in 13 per cent districts but very poor in only 5 per cent districts. The poor to very poor family planning performance in majority of the districts of the country is largely due to poor to very poor capacity of family planning services delivery system to meet the diverse family planning needs of the people.

**Table 3.** Distribution of districts by the level of family planning performance and its components, 2015-16

| Performance level | Modern spacing methods coverage index<br>($c_s$) | | Permanent methods coverage index<br>($c_p$) | | Coverage index<br>© | | Quality index<br>($q$) | | Performance index<br>($p$) | |
| --- | --- | --- | --- | --- | --- | --- | --- | --- | --- | --- |
|  | N | % | N | % | N | % | N | % | N | % |
| Very poor | 163 | 25.5 | 31 | 4.8 | 32 | 5.0 | 338 | 52.8 | 173 | 27.0 |
| Poor | 283 | 44.2 | 67 | 10.5 | 228 | 35.6 | 263 | 41.1 | 373 | 58.3 |
| Average | 146 | 22.8 | 143 | 22.3 | 296 | 46.3 | 39 | 6.1 | 97 | 15.2 |
| Good | 27 | 4.2 | 227 | 35.5 | 82 | 12.8 | 0 | 0.0 | 0 | 0.0 |
| Very good | 1 | 0.2 | 178 | 27.8 | 2 | 0.3 | 0 | 0.0 | 0 | 0.0 |
| Summary measures |  |  |  |  |  |  |  |  |  |  |
| Minimum | 0 |  | 0.031 |  | 0.077 |  | 0 |  | 0.015 |  |
| Q1 | 0.298 |  | 0.667 |  | 0.472 |  | 0.133 |  | 0.289 |  |
| Median | 0.435 |  | 0.808 |  | 0.585 |  | 0.277 |  | 0.403 |  |
| Q3 | 0.584 |  | 0.91 |  | 0.68 |  | 0.448 |  | 0.509 |  |
| Maximum | 0.917 |  | 0.989 |  | 0.949 |  | 0.71 |  | 0.745 |  |
| IQR | 0.285 |  | 0.243 |  | 0.208 |  | 0.314 |  | 0.219 |  |
| CV | 0.402 |  | 0.266 |  | 0.277 |  | 0.599 |  | 0.356 |  |
| N | 640 |  | 640 |  | 640 |  | 640 |  | 640 |  |

Source: Author's calculations

The coverage index *c* is determined by the coverage index of modern spacing methods *c*_*s*_ and coverage index of permanent methods *c*_*p*_. The inter-district variation in the index *c*_*s*_ is different from that in *c*_*p*_. The met demand for modern spacing methods is good or very good in less than 5 per cent districts but poor or very poor in almost 70 per cent districts of the country. By contrast, the met demand for permanent methods is poor or very poor in only about 15 per cent districts but good or very good in more than 63 per cent districts.

District level family planning performance by states/Union Territories is revealing (Table 4). There are eight states/Union Territories where family planning performance is very poor in at least 50 per cent districts of the state. All but one of these states/Union Territories are located in the southern region of the country. On the other hand, there are only seven states where family planning performance is rated as average in at least 50 per cent districts and all but two of these states are located in the northern region. In the remaining states/Union Territories, family planning performance is poor in at least 50 per cent districts. More specifically, there are 19 states/Union Territories, where family planning performance is poor or very poor in all districts whereas in 16 states/Union Territories there is no district where family planning performance is very poor.

**Table 4.** District family planning performance by states/Union Territories.

| Country/State/Union Territory | Family planning performance |  |  | Number of districts |
| --- | --- | --- | --- | --- |
|  | Very poor | Poor | Average |  |
| Andaman & Nicobar Islands | 33.3 | 66.7 | 0 | 3 |
| Andhra Pradesh | 100.0 | 0.0 | 0 | 13 |
| Assam | 6.3 | 56.3 | 37.5 | 16 |
| Arunachal Pradesh | 0.0 | 96.3 | 3.7 | 27 |
| Bihar | 86.8 | 13.2 | 0 | 38 |
| Chandigarh | 0.0 | 0.0 | 100.0 | 1 |
| Chhattisgarh | 22.2 | 77.8 | 0.0 | 18 |
| Dadra & Nagar Haveli | 0.0 | 100.0 | 0.0 | 1 |
| Daman & Diu | 50.0 | 50.0 | 0.0 | 2 |
| Delhi | 0.0 | 33.3 | 66.7 | 9 |
| Goa | 0.0 | 100.0 | 0.0 | 2 |
| Gujarat | 23.1 | 76.9 | 0.0 | 26 |
| Haryana | 0.0 | 38.1 | 61.9 | 21 |
| Himachal Pradesh | 0.0 | 58.3 | 41.7 | 12 |
| Jammu & Kashmir | 0.0 | 40.9 | 59.1 | 22 |
| Jharkhand | 37.5 | 62.5 | 0.0 | 24 |
| Karnataka | 86.7 | 13.3 | 0.0 | 30 |
| Kerala | 78.6 | 21.4 | 0.0 | 14 |
| Lakshadweep | 0.0 | 100.0 | 0.0 | 1 |
| Madhya Pradesh | 40.0 | 60.0 | 0.0 | 50 |
| Maharashtra | 8.6 | 88.6 | 2.9 | 35 |
| Manipur | 11.1 | 88.9 | 0.0 | 9 |
| Meghalaya | 14.3 | 71.4 | 14.3 | 7 |
| Mizoram | 0.0 | 62.5 | 37.5 | 8 |
| Nagaland | 0.0 | 100.0 | 0.0 | 11 |
| Odisha | 0.0 | 90.0 | 10.0 | 30 |
| Puducherry | 75.0 | 25.0 | 0.0 | 4 |
| Punjab | 0.0 | 5.0 | 95.0 | 20 |
| Rajasthan | 0.0 | 97.0 | 3.0 | 33 |
| Sikkim | 0.0 | 0.0 | 100.0 | 4 |
| Telangana | 90.0 | 10.0 | 0.0 | 10 |
| Tamil Nadu | 78.1 | 21.9 | 0.0 | 32 |
| Tripura | 0.0 | 100.0 | 0.0 | 4 |
| Uttar Pradesh | 8.5 | 85.9 | 5.6 | 71 |
| Uttarakhand | 0.0 | 76.9 | 23.1 | 13 |
| West Bengal | 0.0 | 31.6 | 68.4 | 19 |
| India | 27.0 | 57.8 | 15.2 | 640 |

Source: Author's calculations

The district level data also suggests that the simple zero order correlation coefficient between the index *q* and the index *c* is 0.116 which means that inter-district variation in the index *q* or in index *c* accounts for only about 1.3 per cent of the inter-district variation in the index *c* or in index *q*. On the other hand, the simple zero order correlation coefficient between index *p* and index *q* is estimated to be 0.844 while that between index *p* and *c* is estimated to be 0.615. These correlations provide the empirical justification of the index *p* as a measure of family planning performance.

### Classification of Districts

Family planning performance index *p* is the composite of index *q* and index *c* which, in turn, is the composite of indexes *c*_*s*_ and *c*_*p*_. This means that family planning performance in a district can be characterised in terms of indexes *q, c*_*s*_ and *c*_*p*_. The contribution of the three indexes to the index *p* is, however, not additive as different weights are assigned to different indexes in the construction of the index *p*. We have carried out the classification modelling exercise approach (Han, Kamber, Pei, 2012; Tan, Steinbach, Kumar, 2006) to classify districts in terms of index *p* on the basis of indexes *q, c*_*s*_ and *c*_*p*_. The classification and regression tree (CRT) method (Brieman, et al. 1984) was used for the purpose. CRT is a nonparametric recursive partitioning method that divides districts into mutually exclusive clusters in such a way that within-group homogeneity in the index *p* is the maximum. A cluster in which the index *p* is the same for all districts in the cluster is termed as “pure”. If a cluster is not pure than the impurity in the cluster can be measured through the Gini index. If the dependent variable is a categorical one, the method provides cluster-specific distribution of the dependent variable. If the dependent variable is a scale variable, then the method provides estimates of arithmetic mean and standard deviation of the dependent variable within the cluster (Chaurasia, 2018). In the present exercise, the dependent variable, index *p*, is a scale variable. On the other hand, the three explanatory variables, *q, c*_*s*_, *c*_*p*_, are also scale variables. Classification modelling exercise, therefore, provided mean and standard deviation of the inter-district distribution of the index *p* in each cluster. The TREE routine of the SPSS software package has was used for classification modelling.

Results of the classification modelling exercise are summarised in table 5 and the classification tree is depicted in figure 3. The 640 districts of the country, as they existed at the time of 2011 population census, can be grouped into eight mutually exclusive clusters having distinct values of *q, c*_*s*_ and *c*_*p*_ on average. The most important classification variable is the index *q* while the least important classification variable is the index *c*_*p*_. Family planning performance is relatively the best, on average, in 54 (8.4 per cent) districts of cluster 14. These districts are characterised by the index *q* higher than 39.8 per cent, index *c*_*s*_ higher than 60.5 per cent and index *c*_*p*_ less than or equal to 92.5 per cent. On the other hand, family planning performance is relatively the poorest, on average, in 75 (11.7 per cent) districts of cluster 5. These districts are characterised by the index *q* less than or equal to 15.3 per cent and the index *c*_*p*_ more than 92.5 per cent. Family planning performance is also very poor in 68 (10.6 per cent) districts of cluster 7 in which both index *q* and *c*_*s*_ is very low.

**Table 5.**
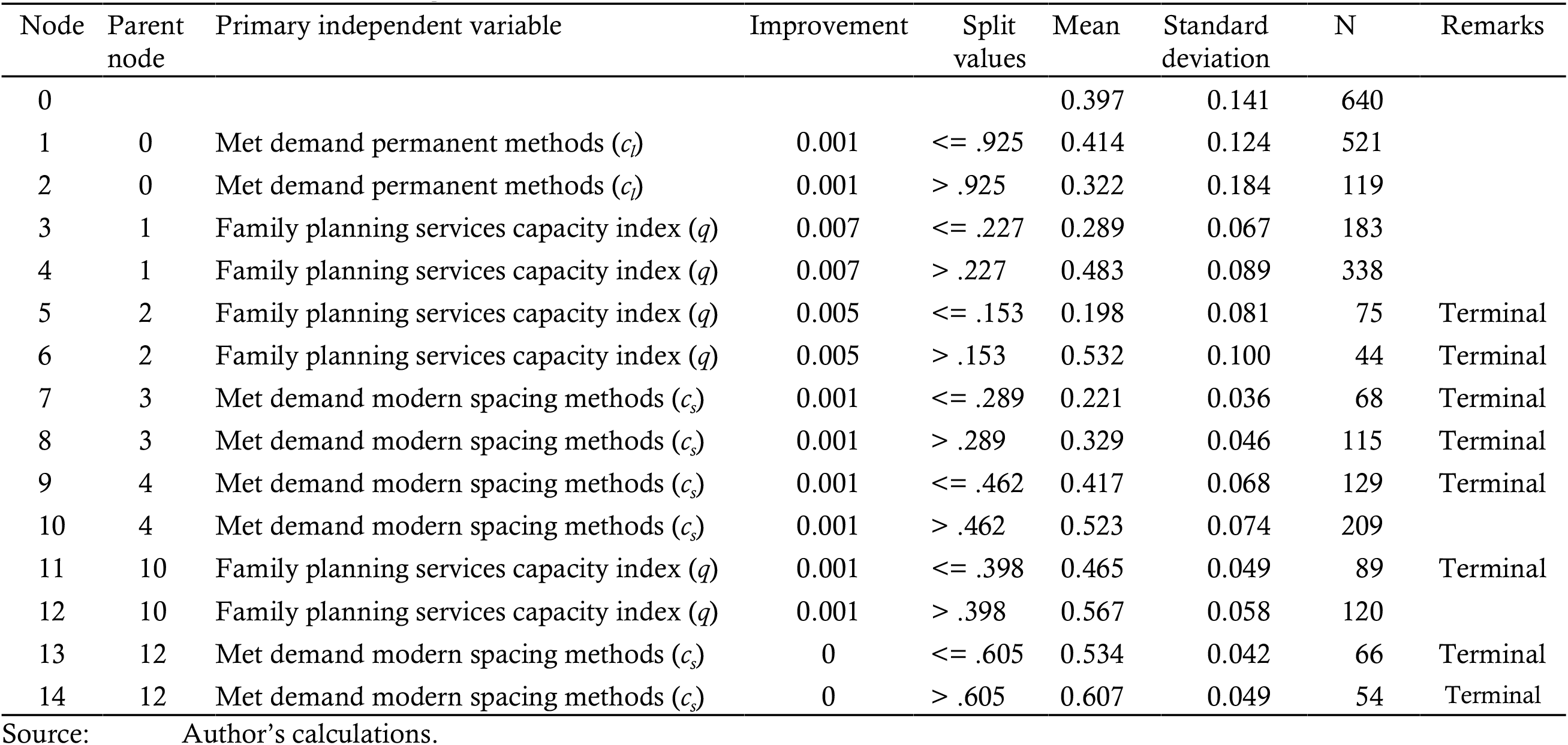
Results of the classification modelling exercise.

**Figure 1.**
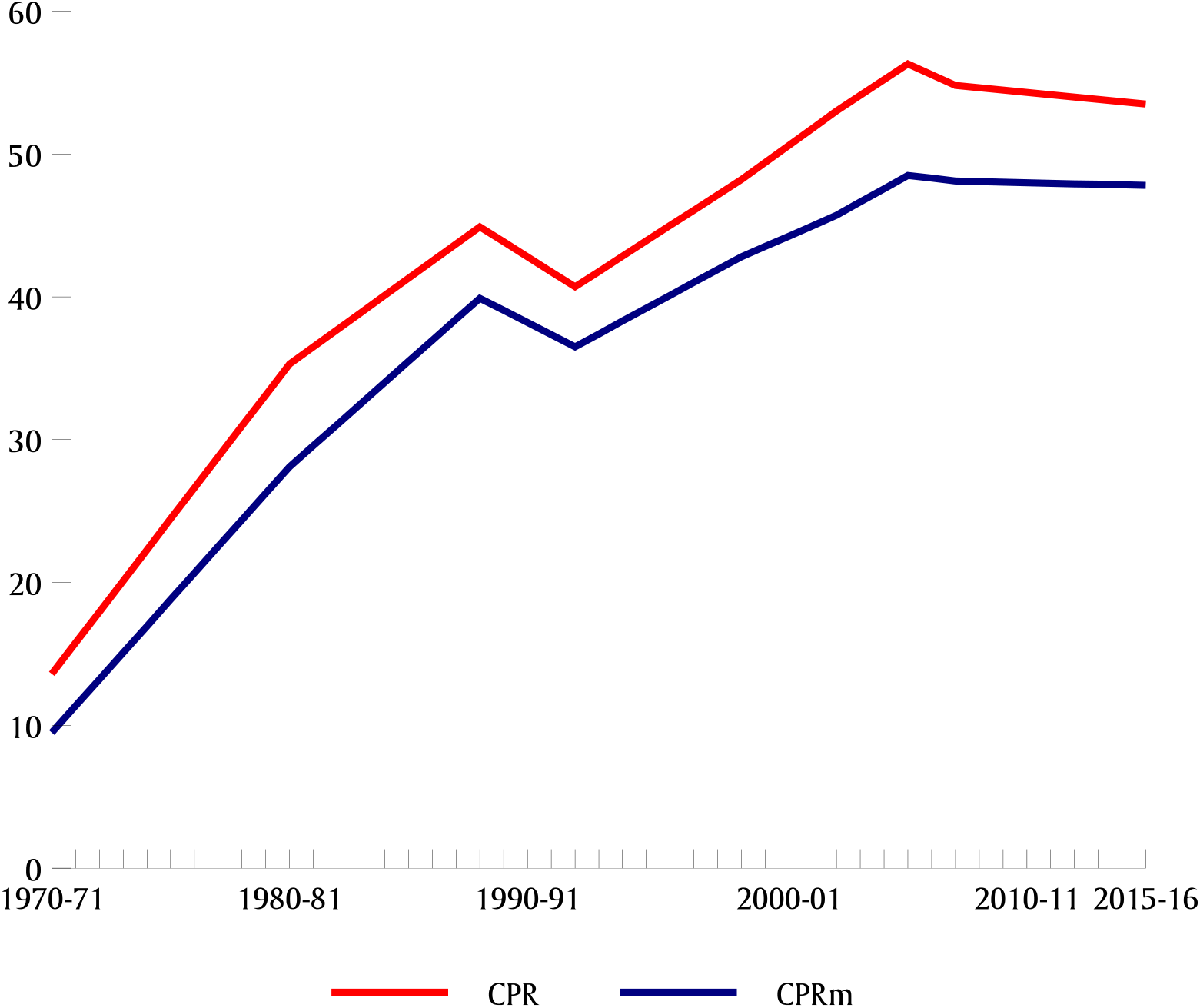
Trend in *CPR* and *CPRm* in India

**Figure 2.**
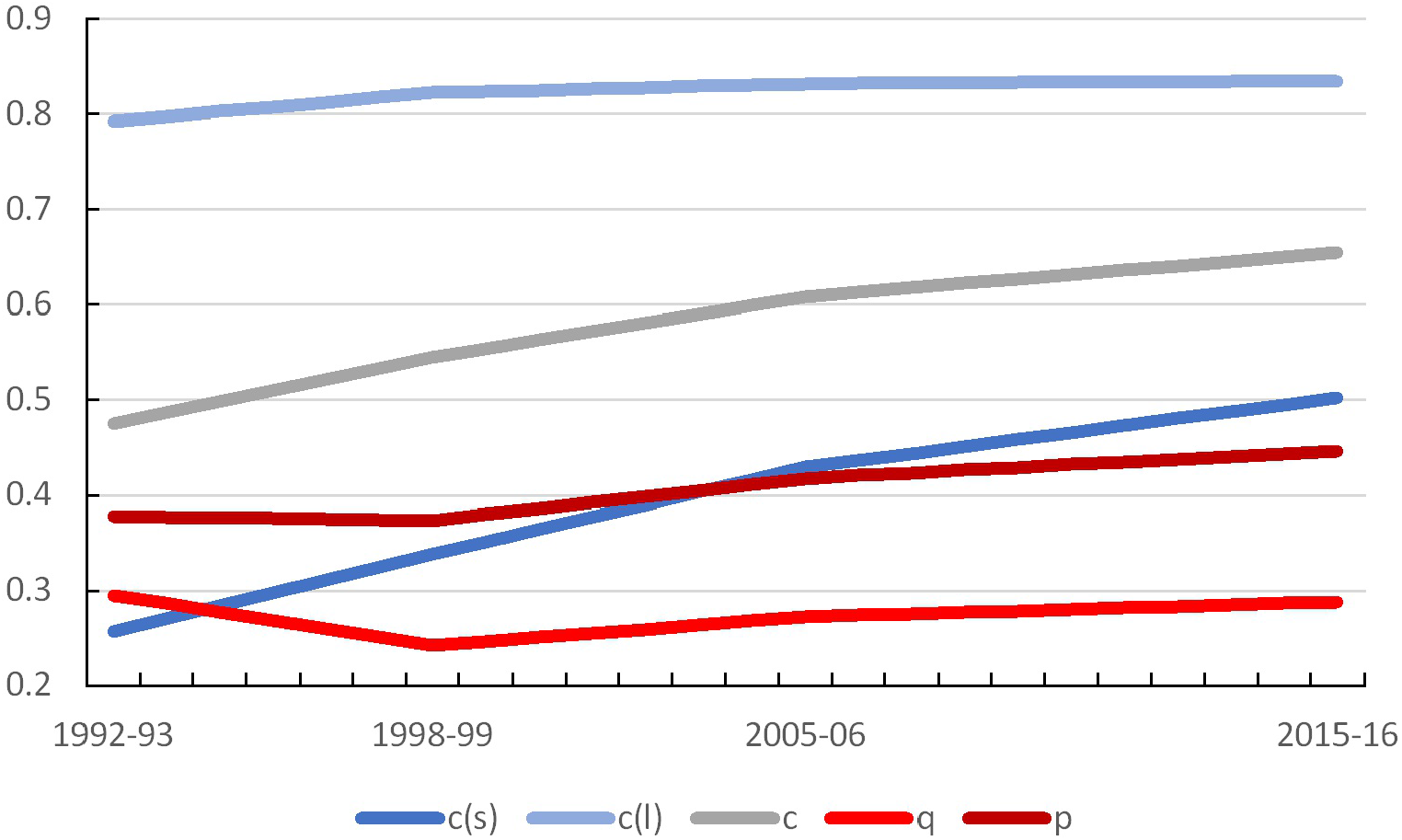
Trend in family planning performance indicators in India 1992-93 through 2015-16

**Figure 3.**
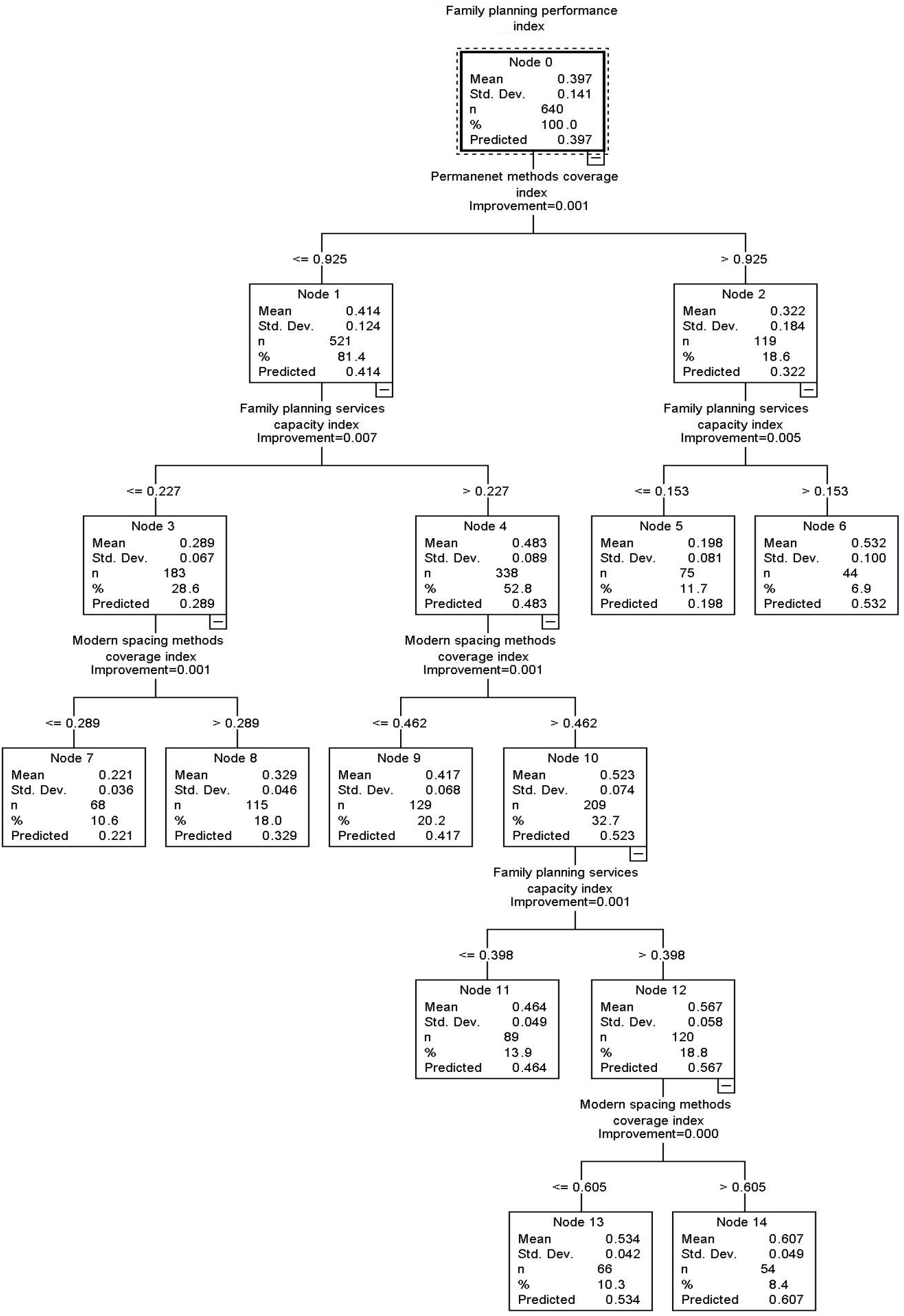
Classification Tree

Table 6 presents the distribution of districts in different clusters by the level of the index *p*. In cluster 14, family planning performance is average (0.550≤*p*<0.750) in more than 90 per cent districts. There are only five districts in this cluster where family planning performance is poor (*p*<0.550). By contrast, family planning performance is very poor (*p*<0.300) in more than 97 per cent districts belonging to cluster 7 and almost 90 per cent districts of cluster 5. There are only two districts in cluster 7 and 8 districts in cluster 5 where family planning performance is poor (0.300≤*p*<0.550). There is no district in clusters 6, 11, 13 and 14 where family planning performance is very poor (*p*<0.300). On the other hand, there is no district in clusters 5, 7 and 8 where family planning performance is either poor or average. There is no district where family planning performance is either good or very good.

**Table 6.** Distribution of districts by family planning performance index in different clusters identified through classification modelling exercise

|  | Cluster |  |  |  |  |  |  |  |
| --- | --- | --- | --- | --- | --- | --- | --- | --- |
|  | 5 | 6 | 7 | 8 | 9 | 11 | 13 | 14 |
| *Defining characteristics* |  |  |  |  |  |  |  |  |
| $q$ | $\leq 0.153$ | $> 0.153$ | $\leq 0.227$ | $\leq 0.227$ | $> 0.227$ | $> 0.227 \leq 0.398$ | $> 0.398$ | $> 0.398$ |
| $c_s$ | | | $\leq 0.289$ | $> 0.289$ | $\leq 0.462$ | $> 0.462$ | $> 0.462 \leq 0.605$ | $> 0.605$ |
| $c_p$ | $> 0.952$ | $> 0.952$ | $\leq 0.925$ | $\leq 0.925$ | $\leq 0.925$ | $\leq 0.925$ | $\leq 0.925$ | $\leq 0.925$ |
| *Frequency* |  |  |  |  |  |  |  |  |
| Very poor | 89.3 | 0.0 | 97.1 | 27.8 | 6.2 | 0.0 | 0.0 | 0.0 |
| Poor | 10.7 | 45.5 | 2.9 | 72.2 | 93.0 | 97.8 | 68.2 | 9.3 |
| Average | 0.0 | 54.5 | 0.0 | 0.0 | 0.8 | 2.2 | 31.8 | 90.7 |
| *Summary measures* |  |  |  |  |  |  |  |  |
| Mean | 0.198 | 0.532 | 0.221 | 0.329 | 0.417 | 0.464 | 0.534 | 0.607 |
| SD | 0.081 | 0.100 | 0.036 | 0.046 | 0.068 | 0.049 | 0.042 | 0.049 |
| CV | 0.409 | 0.188 | 0.163 | 0.140 | 0.163 | 0.106 | 0.079 | 0.081 |
| Minimum | 0.015 | 0.359 | 0.138 | 0.231 | 0.236 | 0.326 | 0.434 | 0.461 |
| Median | 0.200 | 0.568 | 0.220 | 0.320 | 0.419 | 0.470 | 0.535 | 0.599 |
| Maximum | 0.358 | 0.677 | 0.309 | 0.435 | 0.551 | 0.580 | 0.671 | 0.745 |
| IQR | 0.126 | 0.174 | 0.050 | 0.066 | 0.090 | 0.060 | 0.048 | 0.037 |
| N | 75 | 44 | 68 | 115 | 129 | 89 | 66 | 54 |

Source: Author's calculations

Regional pattern of district family planning performance is also evident from the classification modelling exercise (Table 7; Figure 4). More than 63 per cent districts included in cluster 14, are located in the northern region of the country while another 35 per cent are located in the eastern region. There is only one district in this cluster which is located in the central region whereas there is no district in this cluster which is located in either western or southern regions of the country. On the other hand, 80 per cent of districts in cluster 5 are located in the southern region and the remaining ones are located in central and western regions. There is no district in this cluster which is located in northern and eastern regions of the country. The strong regional difference in family planning performance, as revealed through the classification modelling exercise suggests that the orientation of family planning services delivery system is essentially different in different regions of the country. Since family planning services delivery in the country is largely contingent upon the official family planning efforts, this implies that there are regional differences in the basic orientation of the family planning services delivery system in the country which has an impact on family planning performance.

**Table 7.**
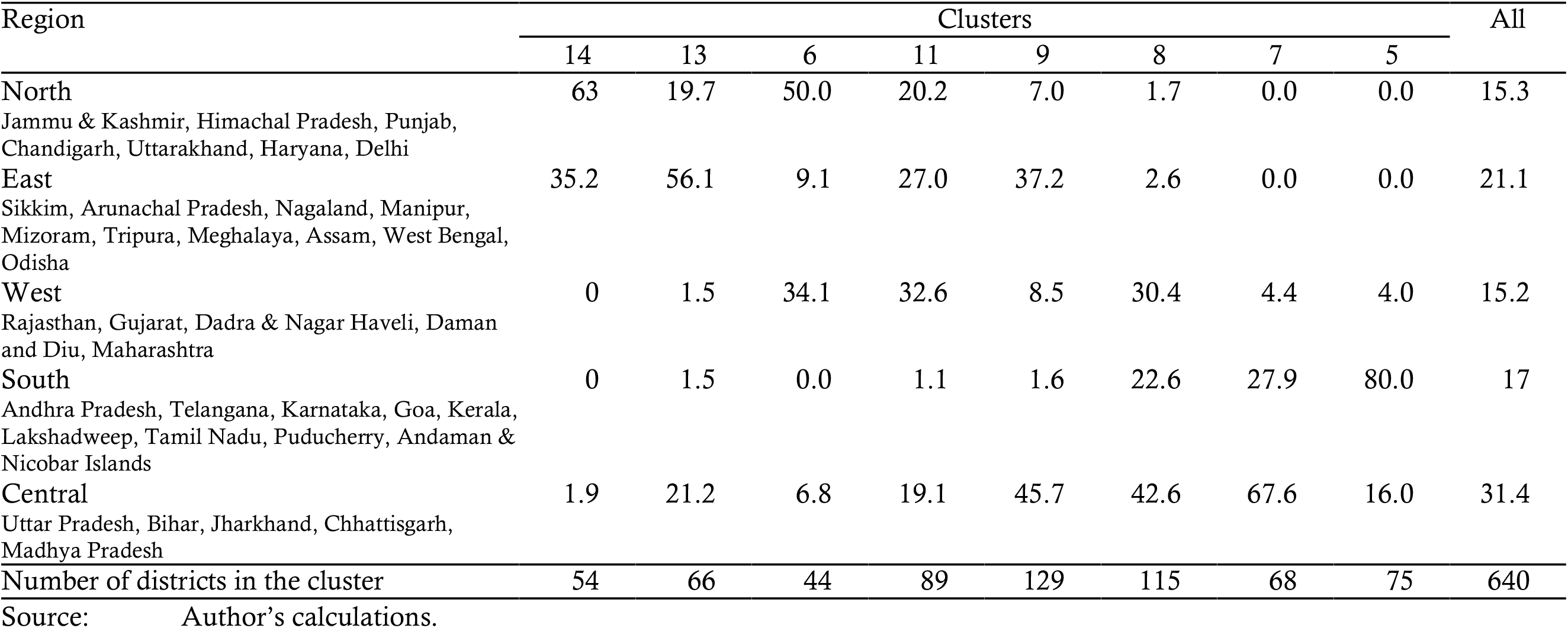
Regional patterns of family planning performance in India, 2015-16 (Proportion of districts in different clusters by region)

| Region | Clusters |  |  |  |  |  |  |  | All |
| --- | --- | --- | --- | --- | --- | --- | --- | --- | --- |
|  | 14 | 13 | 6 | 11 | 9 | 8 | 7 | 5 |  |
| North | 63 | 19.7 | 50.0 | 20.2 | 7.0 | 1.7 | 0.0 | 0.0 | 15.3 |
| Jammu & Kashmir, Himachal Pradesh, Punjab, Chandigarh, Uttarakhand, Haryana, Delhi |  |  |  |  |  |  |  |  |  |
| East | 35.2 | 56.1 | 9.1 | 27.0 | 37.2 | 2.6 | 0.0 | 0.0 | 21.1 |
| Sikkim, Arunachal Pradesh, Nagaland, Manipur, Mizoram, Tripura, Meghalaya, Assam, West Bengal, Odisha |  |  |  |  |  |  |  |  |  |
| West | 0 | 1.5 | 34.1 | 32.6 | 8.5 | 30.4 | 4.4 | 4.0 | 15.2 |
| Rajasthan, Gujarat, Dadra & Nagar Haveli, Daman and Diu, Maharashtra |  |  |  |  |  |  |  |  |  |
| South | 0 | 1.5 | 0.0 | 1.1 | 1.6 | 22.6 | 27.9 | 80.0 | 17 |
| Andhra Pradesh, Telangana, Karnataka, Goa, Kerala, Lakshadweep, Tamil Nadu, Puducherry, Andaman & Nicobar Islands |  |  |  |  |  |  |  |  |  |
| Central | 1.9 | 21.2 | 6.8 | 19.1 | 45.7 | 42.6 | 67.6 | 16.0 | 31.4 |
| Uttar Pradesh, Bihar, Jharkhand, Chhattisgarh, Madhya Pradesh |  |  |  |  |  |  |  |  |  |
| Number of districts in the cluster | 54 | 66 | 44 | 89 | 129 | 115 | 68 | 75 | 640 |

Source: Author's calculations.

**Figure 4.**
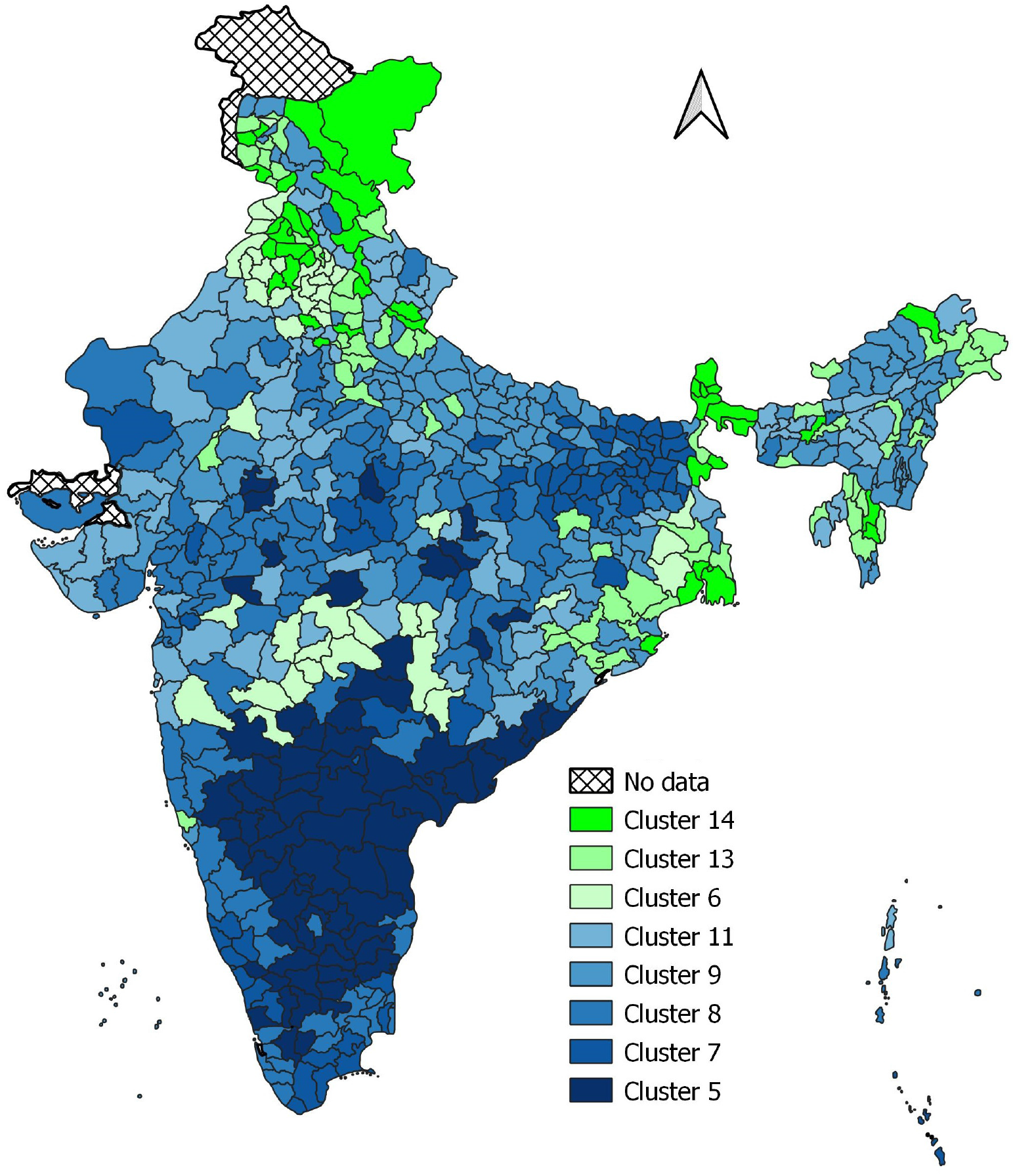
Cluster patterns

## Discussions and Conclusions

This paper employs, probably and so obviously for the first time, a composite index to measure family planning performance. The proposed composite performance index may serve as the basis to convey summary information about the performance of family planning efforts in meeting family planning needs of the people and may signal priorities in improving the delivery of family planning services. The index offers a more rounded assessment of family planning performance and presents the ‘big picture’ in a simple and convincing manner that appeals to policy makers, programme managers and even to the common people. Another advantage of the index is that it may be calculated from the already available data.

The application of the composite performance index to India suggests that India’s family planning performance remains poor despite sustained official efforts spanning almost seven decades. Delivery of family planning services in India has almost entirely been a prerogative of the official family planning efforts. Poor family planning performance of the country, therefore, reflects, the poor performance of the organised family planning efforts in meeting the family planning needs of the people. The analysis also reveals very strong, inter-district, variation in family planning performance within the country. At the district level, family planning performance appears to be significantly influenced by district-specific factors both endogenous and exogenous to the district family planning services delivery system. Endogenous factors are related to the organisation of family planning services. Very little is currently known about the organisation of district family planning services despite the fact that the capacity of family planning services delivery system in meeting diverse family planning needs of the people is largely determined by the way family planning services are organised at the district level. On the other hand, exogenous factors like rural-urban distribution of the population, level of education, religious and social class composition of the population, also influence family planning performance. Although, exogenous factors exert a strong impact on family planning performance, yet, this impact can be minimised by improving the organisational efficiency of the family planning services delivery system.

In conclusion, the present analysis emphasises the need of a comprehensive reinvigoration of family planning services delivery system, especially, official family planning efforts so as to effectively meet the diverse family planning needs of the people. The official family planning efforts in India appear to be organised under the narrow perspective of fertility reduction through birth limitation. This perspective needs to be relaced by a family building approach which emphasises proper spacing between marriage and first birth as well as between successive births. The composite family planning performance index, proposed in this paper, may serve as the basis for inducing such strategic shift in the delivery of family planning services, especially official family planning services. This strategic shift in the organisation of family planning services is also necessary because an increasing number of states and Union Territories of the country has now achieved replacement fertility. In these states/Union Territories, birth limitation-based approach of family planning services is now largely irrelevant. There is already evidence that a major share of future population growth in India will be the result of the momentum built-in the young age structure of the population (Chaurasia and Gulati, 2007; Chaurasia 2016). Momentum effects of population growth can be minimised only by either lowering the completed fertility below the replacement level or by increasing the mean age at child bearing through birth planning - delaying the entry into marital union and increasing spacing between births (Bongaarts, 1994). Shifting the focus from birth limitation to birth planning will also contribute to accelerating the reduction in infant, child and maternal mortality. It is, therefore, important that family planning is treated as a development strategy and integrated in the development agenda of the country rather than an intervention to limit births and reduce fertility. At the policy level, the need for such a shift has been recognised in India’s ‘Vision FP 2020’ document (Government of India, 2014). The challenge, however, remains to translate this vision into practice.

## Data Availability

All data used in the analysis are available in public domain.

